# ECONOMIC AND HEALTH IMPACTS OF BOVINE TUBERCULOSIS ON RURAL ZAMBIAN COMMUNITIES

**DOI:** 10.1101/2025.10.15.25336428

**Authors:** Anthony Phiri, Mildred Zulu, Maxwell Phiri, Mulenga Malata, Sydney Kalenga, Sydney Malama

## Abstract

**Background:** Bovine Tuberculosis (bTB) is a persistent and significant challenge for cattle farmers in Zambia, especially among rural farmers. As livestock farming constitutes a crucial component of the country’s agricultural economy, the prevalence of bTB threatens not only animal health and productivity but also poses serious risks to human health, food safety, and economic stability within rural communities.

**Methods:** A mixed-methods study was conducted in Lundazi and Monze districts of Zambia, between December 2021 and June 2022, combining a cross-sectional survey of 280 respondents with qualitative insights from five focus groups and five key informant interviews. Data analysis was done using the R software for quantitative data and NVivo for qualitative data.

**Results:** Our study indicates that cows infected with Bovine tuberculosis (bTB) experience an average decline in milk production of approximately 3.75 liters, which translates to a substantial economic loss of around ZMW 10.00 per cow, based on an average milk price of ZMW 8.00 per liter (equivalent to an exchange rate of ZMW 18.17 per USD).

Study findings also revealed that bTB infection significantly reduced monthly income from livestock farming, with a strong association between bTB impact and decreased income.

Further, among the education levels, only the primary education level was significantly impacted. Other education levels were not significantly impacted by bTB; however, the odds of being impacted by bTB were lower compared to other education levels. There were no significant differences in the impact of bTB on the type of occupation.

Our qualitative findings indicate that rural elderly individuals, specifically those in the 40-50 and above age category, bear a disproportionate burden of bTB’s impact on public health, with a significantly higher likelihood of experiencing adverse effects.

**Conclusions:** Our study reveals true evidence of the significant impact of bTB on rural cattle farming, with critical implications for policy and practice. It highlights the need for appropriate interventions to address the disproportionate burden of bTB among vulnerable populations, such as older people and those with primary education. Prioritizing bTB control and prevention can minimize the economic and health impacts of the disease and promote more sustainable and resilient livestock farming systems.

## INTRODUCTION

Bovine tuberculosis (bTB) is a significant public health concern in rural Zambia, particularly in remote rural areas where livestock farming is the most important part of the agricultural economy. The disease, caused by Mycobacterium bovis, can be easily transmitted to humans through the consumption of contaminated animal products or direct contact with infected animals. Zambia is among the countries with the maximum burden of tuberculosis, with an estimated 60,000 people contracting TB annually, and approximately 15,000 succumbing to the disease every year [1].

The impact of bTB on human health in Zambia is substantial, with studies indicating that the disease disproportionately affects mostly the rural vulnerable populations, including those with limited access to healthcare services and those living in poverty.

Bovine tuberculosis triggers substantial socioeconomic costs, which are estimated at $3 billion yearly, as a result of reduced cattle productivity, milk production, and trade restrictions[1]. Reports in Indian revealed the prevalence of bTB, which is estimated at around 7.3%, affecting approximately 21.8 million cattle [2] [1]

Bovine tuberculosis is a zoonotic disease that is transmissible to humans through the consumption of undercooked meat, unpasteurized dairy products, and handling with bare hands at the abattoir. In the African region, about 82% of people and 85% of cattle reside in regions where bTB is prevalent[2].

Bovine tuberculosis is a significant public health concern in Zambia, more especially among the rural livestock farmers who do not practice hygiene. It was revealed that rural livestock farmers in Zambia have poor awareness of bTB transmission, with 75.3% of males and 70.3% of females expressing inadequate knowledge [1][17].

In Zambia, the prevalence of bTB has been associated with Kafue Lechwe, which act as reservoir hosts and share grazing fields with domestic animals, mostly cattle [2]. This reveals the need for an intensive approach to control and prevent the transmission of bTB, including addressing the disease in both animals and humans.

Despite the significance of bTB being a public health concern, there is limited information on the impact of the disease on the rural human health population in Zambia. Numerous studies have been conducted, focusing primarily on the prevalence of bTB in cattle, with minimal attention paid to the human health implications and challenges. This continuous lack of information makes it challenging to develop effective public health strategies to control and prevent the spread of bTB [3].

Therefore, this study aimed to investigate the impact of bTB on public health in the rural population of Zambia, with a focus on identifying high-risk demographic groups and understanding the factors associated with bTB transmission. By understanding the impact of bTB on human health, policymakers and healthcare professionals can develop targeted interventions and public health strategies to mitigate the burden of the disease. Furthermore, this study contributes to a better understanding of the impact of bTB on public health in Zambia and informs evidence-based policies to protect the less privileged people in the population.

The study’s findings also have major implications for public health policy and practice in Zambia. By identifying the demographic groups most affected by bTB, policymakers and healthcare professionals can therefore develop tailored approaches to disease control and prevention. This may include targeted education and awareness campaigns, improved access to healthcare services, and enhanced surveillance and monitoring of bTB cases [2][4].

Currently in Zambia, the social determinants of tuberculosis, including poverty, lack of access to healthcare services, and poor living conditions, have been shown to contribute to the high burden of TB in the rural communities [3]. Additionally, the association between TB and HIV co-infection is also an imperative concern, with studies indicating that individuals with HIV are more likely to develop active TB [3]. Understanding the social determinants of bTB and their association with TB/HIV co-infection is crucial for developing effective public health strategies to control and prevent the spread of the disease.

The study’s findings also inform the development of gender-tailored tuberculosis health promotion and case-finding strategies. Studies have shown that men and women have different experiences and challenges in accessing TB diagnosis and treatment, and that gender-sensitive approaches are needed to address these disparities [5].

In conclusion, this study aimed to contribute to a better understanding of the impact of bTB on public health in Zambia and inform the development of evidence-based policies to protect vulnerable populations. By identifying the high-risk demographic groups and understanding the factors associated with bTB transmission, policymakers and healthcare professionals can develop targeted interventions and public health strategies to mitigate the burden of the disease.

### Null and Alternative Hypotheses

1. Null Hypothesis (H0): There is no significant association between bTB economic and public health outcomes among rural Livestock farmers in Lundazi and Monze districts.
2. Alternative Hypothesis(H1): There is a significant association between bTB economic and public health outcomes among rural Livestock farmers in Lundazi and Monze districts, with increased risk of infection and adverse health outcomes among those with direct contact with infected animals.

## METHOD

### Study Sites

This research reveals critical public health concerns in districts where humans and animals coexist closely [6], and where the community practices the consumption of unpasteurized milk [7]. The study was conducted in Lundazi and Monze districts of Zambia (Figure 1). Lundazi is located in the eastern part of Zambia (12.2849° S, 33.1745° E), while Monze is located in the southern part of Zambia (16.2803° S, 27.4733° E).[8]. These areas boast of substantial cattle populations and frequent human-animal interactions [9]

**Figure 1:**
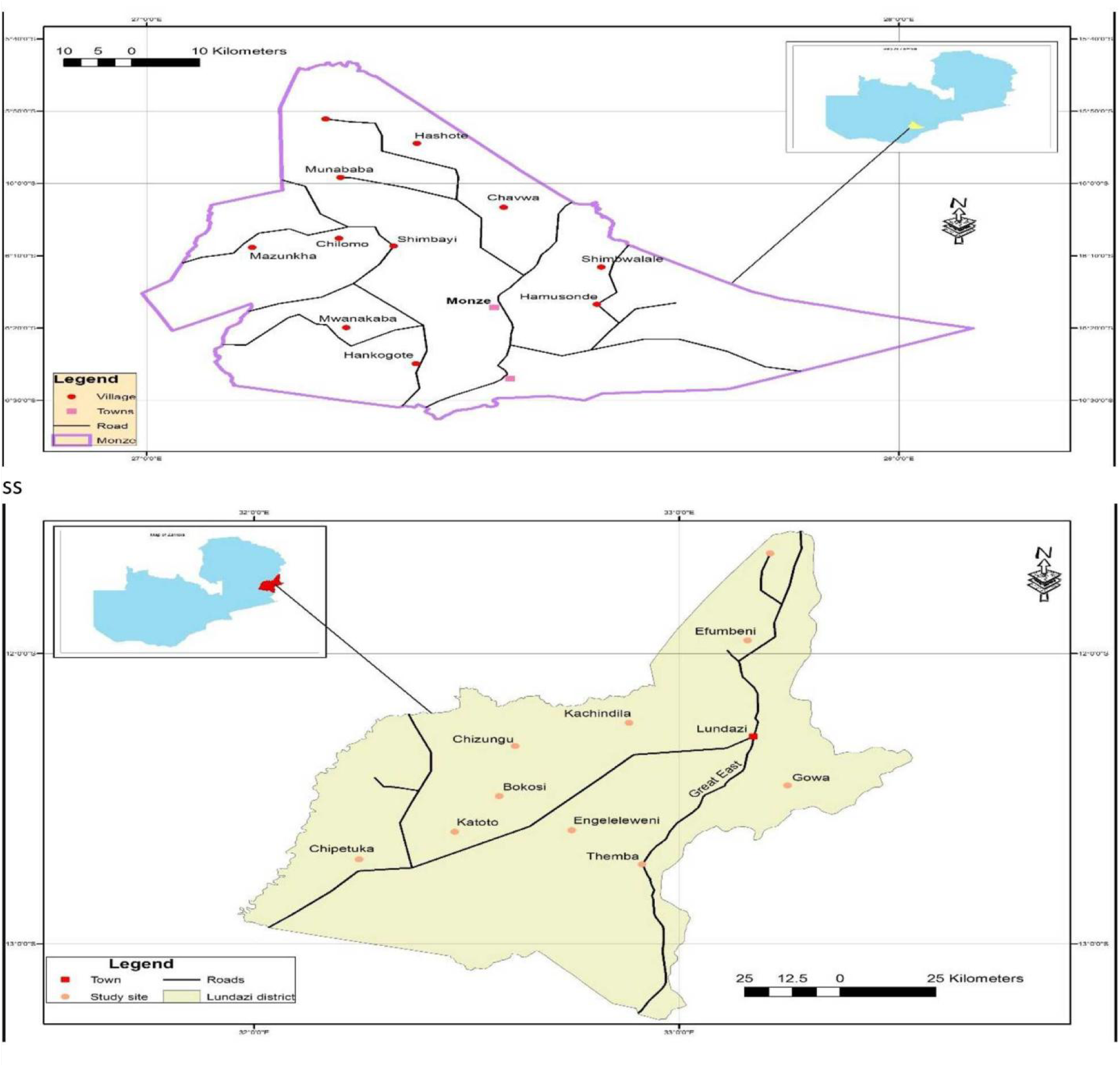
Map of Lundazi and Monze Districts based on the LayerStack process (developed by authors) Reprinted from [Zambia_Mosaic_250Karc1950_ddecw] under a CC BY licence, with permission from the Surveyor General, Government Republic of Zambia, original copyright, [1974]

### Sample Design

According to Kothari [10] [11], a research design informs decisions concerning a research study and the arrangement of conditions for the collection and analysis of data to combine relevance to the research. To achieve the set objective, the research used a mixed-methods cross-sectional design. A key feature of mixed-methods research is its methodological pluralism or eclecticism, which frequently results in superior research compared to mono-method research [11]. Additionally, the cross-sectional design was chosen because collecting data at a single point in time is economical in terms of time, financial resources, and the nature of the study objectives [10]. (That is, it used both quantitative and qualitative methods) to collect data.

### Study Population and Sampling Strategy

The population of interest was cattle farmers in Lundazi and Monze districts. A random sampling strategy was used to select 280 respondents for the cross-sectional survey. Focus groups (n=5) and key informant interviews (n=5) were conducted to gather qualitative data.

### Data Collection Methods and Challenges Interview Duration

The researcher conducted interviews for approximately 60 minutes, which is within the very much recommended time frame for in-depth interviews [10].

To ensure accuracy and validity, the researcher employed multiple methods. (a)Note-taking in a field notebook and (b) audio recording with participant consent [11] (c) Semi-structured interview guides, (d) Participant observation [12][13][15]

### Quantitative Data Collection Methods

The researcher conducted a Cross-sectional survey of 280 respondents (208 livestock farmers) [17]

### Data Analysis Tools

Analysis was analyzed through (a) Thematic analysis (qualitative) [13][14], (b) R software (quantitative) [16], (c) NVivo software for data management and coding (qualitative) [15]

### Challenges

The researcher encountered the following challenges during data collection.(a) Time and resource constraints, (b) adverse weather,(c) inaccessible roads,(e) farmers’ unavailability during rainy season [12] |

### Triangulation and Validation

To make sure that the validity and reliability of the findings is enhanced, analytical frameworks were used to cross-check narratives from key informants with those of focus group participants, which revealed divergent or supporting views. Illustrative quotations that represented the themes were used to support the results, providing a rich and contextualized understanding of the research phenomenon.

The University of Zambia Biomedical Research Ethics Committee (UNZABREC) granted ethical approval for this study (reference number 2102-2021). We also secured authorization from the Provincial Veterinary Officers in the two provinces to collect animal-related data

## RESULTS

### Quantitative

Loss of milk and meat production due to strong cough (bTB) at the household level (ZMW 18.17 per USD)

It was observed that a diseased cow (strong cough) will incur a reduction in milk production of an average of close to 3.75 liters. The cost of 1 liter of milk was sold at an average price of ZWK8.00. Therefore, 5 liters by ZWK8 will give ZWK40.00. Alternatively, low production of milk yields at 3.75 liters by ZWK8 will give ZWK 30. The income is reduced by a ZWK10 (Table 1)

**Table 1.** Estimated Cattle milk production diseased and non-diseased category, animal, production (L), cost (1L/ZWK), overall cost (ZWK), economic loss (ZWK). Source: Own preparation based on data from the Lundazi Market as of 21 February 2022. and Bweengwe small market on 17^th^ June 2022.

| Measure | Animal | Production (L) | Cost(1L/K) | Overall Cost K | Economic loss(K) |
| --- | --- | --- | --- | --- | --- |
| Non-Diseased | 1 | 5 | 8 | 40.00 |  |
| Diseased | 1 | 3.75 | 8 | 30.00 |  |
|  |  |  |  |  | **10.00** |

Quantitative and qualitative data were taken from Lundazi and Bweengwe’s small market.

### The Disease affects Income levels

The study further determined the impact of bTB on the monthly income generated from rural cattle farming. There was a significantly high impact of bTB on monthly income (p< 0.001, OR = 0.99, CI = 0.997-0.998). For every 0.99 odds of a decrease in monthly income, the impact from bTB increased, an indication of a reduction in monthly income resulting from the high impact of bTB. (Figure 2 A and B)

**Figure 2.**
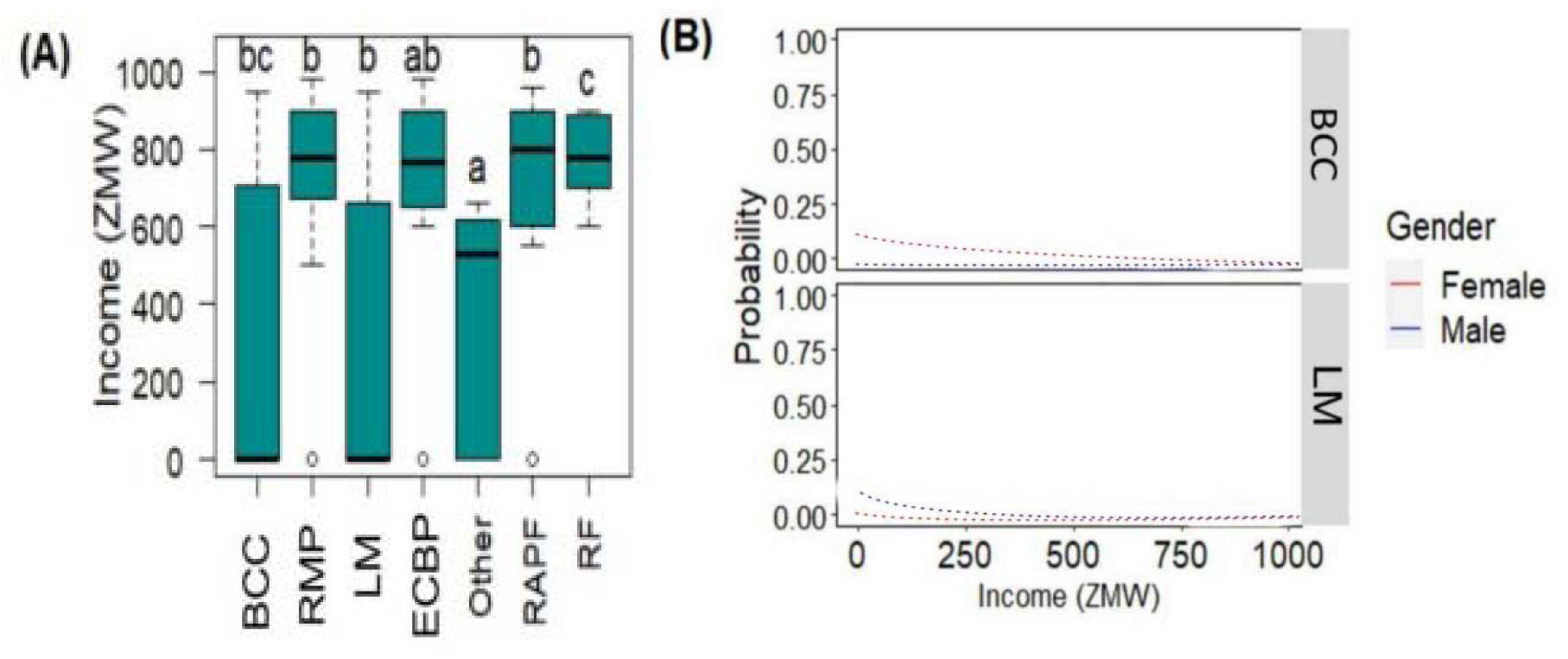
The impact of bTB (BCC: Beef Carcass Condemnation, RMP: Reduced Meat Production, LM: Low Milk production, ECBP: Exchange Cattle for Bridal Price, Other: unspecified reasons, RAPF: Reduced Animal Power for Farming, and RF: Reduced Fertility) on monthly income ZMW (A). Analyzed from a generalized linear model with lower case letter showing significant differences at p < 0.05. The change in predicted mean probabilities of income attributed to bTB impact factors of BCC and LM on Gender (B) Among the education levels, only the primary education level was significantly impacted by bTB (p = 0.002, CI = 1.60-8.23, OR = 3.55). Other education levels were not significantly impacted by bTB (Table 2); however, the odds of being impacted by bTB were 1.24 times lower compared to other education levels. There were no significant differences in the impact of bTB on the type of occupation (p> 0.05).

**Table 2.** The impact of bTB on education levels in Zambia. The confidence intervals are indicated at 95% and the significant differences are considered at *p*< 0.05 using ordinal logistic regression.

| Education level | Odds Ratio | Confidence Intervals | *p*-value |
| --- | --- | --- | --- |
| Primary | **3.55** | **1.60-8.23** | **0.002** |
| Secondary | 1.28 | 0.53 - 3.21 | 0.584 |
| College | 2.08 | 0.95 - 4.74 | 0.071 |
| University | 1.32 | 0.43 – 4.01 | 0.615 |
| Postgraduate | 1.24 | 0.14 – 7.69 | 0.824 |

Source: Authors, field data

### Qualitative

Four main themes were yielded from the focus group discussions and interviews as follows;

- Economic Burden due to bTB
- Vulnerability of elderly people in the community
- Knowledge and Awareness Gaps Analysis
- Challenges in Controlling bTB and Prevention

### Economic Burden due to bTB

It is a culture among most cattle farmers to generate income from their animals. However, when an animal has been affected by bTB. The production of milk and meat decreases drastically.

The findings indicate that the incomes of the respondents were very much affected due to the diseased animals, which could not produce the desired volume of milk.

> *“I have three animals suffering from a strong cough, and the production of milk has since reduced. Initially, 1(one) animal used to produce close to 5 liters; unfortunately, to date, 1(one) animal produces only close to 3 liters of milk. Table 1“*
>
> *[Old age Male in Lundazi]*
>
> *‘Most of my people in my village had experienced carcass condemnation at the abattoir, they would want to sell their animals, unfortunately, some parts like the lungs, the liver, and other organs of the cow”*
>
> *[Village Headman in Lundazi]*
>
> *“I took my animal to an abattoir to be sold so that I can generate money for my firstborn child’s school fees. However, at an abattoir, some parts like the lungs and organs of the digestive system were condemned, such that I could not gain enough money as I expected.”*
>
> [Old age Male in Monze].
>
> *“I could not send my two children to high school because the money that I used to generate through the sales of milk had additionally reduced, and we could not afford three meals a day. As we used to, he lasts seven years. After a good sale of milk at our local market”*
>
> *[Middle-aged Female in Monze]*

### Vulnerability of elderly people in the community

Our findings also revealed that elderly people who have been in the rearing and selling of milk and have close contact with abattoir workers have been deeply affected by the disease.

> *“I have been handling and selling meat, milk, and meat products since I was young, and I have always consumed unpasteurized milk. However, two years ago, I fell ill with a fever and was referred to the main hospital in Lusaka. After some tests, I was diagnosed with tuberculosis. The doctor asked about my occupation, and I told him I’m a cattle farmer who sells milk. He advised me, at 63, to let my children or younger people handle the milk sales, as my immune system is weak due to age and I’m more susceptible to zoonotic diseases like TB.”*
>
> *[Old age Male in Lundazi]*
>
> *“I grew up feeding on unpasteurized milk and on meat that was not fully cooked. I would consume meat that has not been fully prepared from beer-drinking places.”*
>
> *[Old age Female in Monze]*

### Knowledge and Awareness Gaps Analysis

Some respondents were not aware that Bovine Tuberculosis affects human health as well. They thought bTB is a disease that belongs to animals and cannot be transmitted to human beings.

> *‘’I now come to realize that bovine tuberculosis (bTB) can be easily transmitted to humans through close contact with infected animals, handling meat and meat products, consuming undercooked meat, and drinking unpasteurized milk.”*
>
> *[Middle-aged Male in Monze]*
>
> *” In my village, my people like the traditional way of treating diseases, which may contribute to the spread of bTB, and potential opportunities for modifying these practices.”*
>
> *[Village Headman in Lundazi]*
>
> *“All along, I have been using traditional medicine to treat my animals when they are sick. I did not know that feverishness, loss of weight, loss of appetite, and prolonged cough in animals are the clinical signs of zoonotic disease like bTB”.*
>
> *[Middle-aged Male in Lundazi]*
>
> *“I used to think that the only way to control and eliminate this disease, which affects the low production of milk and meat in our animals, is to have animals vaccinated.”*
>
> *[Old age Female in Lundazi]*

Some respondents also highlight limited information about bTB and a lack of resources :

> *“Regrettably... ... ... ... I have never received any piece of guidance or information about bTB from either human doctors or veterinary Doctors, specifically on how to effectively treat or control the disease. As a result, when my animals fall ill, I have been forced to rely on my limited knowledge to administer treatment without any professional prescription or guidance.*
>
> *Unknowingly to me, the medications I administer to my animals may be causing more harm than good, potentially leading to long-term suffering and complications for the animals.”*
>
> *[Old age Male in Monze]*
>
> *‘’In my village, we do not seem to have access to testing or diagnostic services for bTB, hence there is an impact of the disease’’*
>
> *[Middle-aged Male in Lundazi]*

### Challenges in Controlling bTB and Prevention

Our qualitative data revealed potential challenges in controlling and preventing bTB among cattle farmers in Zambia, including limited access to resources, knowledge, and infrastructure. Therefore, by understanding these challenges, targeted interventions and support programs can be developed to address these issues and improve bTB control and prevention. Below are some of the responses:

> *“I have never seen an effort by the government in terms of funding for bTB control programs; however, much of the funding has been channeled towards the fight against malaria.”*
>
> *[Village Headman in Lundazi]*
>
> *“I have been rearing cattle for more than 40 years; however, I am not aware of the best practices for handling milk, meat, and meat products.”*
>
> *[Old age Female in Lundazi]*
>
> *“We have been consuming unpasteurized milk and undercooked meat; we do not see why we should change our lifestyle now. “*
>
> *[Old age Male in Monze]*
>
> *“Some farmers are very skeptical about the existence risk of bTB in humans, hence they don’t take precautions.”*
>
> *[Village Headman in Monze]*
>
> *“In our villages, there is a serious lack of proper facilities, which makes it hard to maintain hygiene and prevent the spread of bTB.”*
>
> *(Middle-aged Male in Lundazi)*

## DISCUSSION

Bovine Tuberculosis (bTB) has a significant economic impact on milk production and meat yield in Zambia. Findings from the milk sales markets in the study area revealed household loss due to the impact of bTB. Milk sales information from the milk markets showed a reduction in milk production, with diseased cows experiencing a significant reduction in milk production, averaging approximately 3.75 liters per cow [17] [18] [19]. In Comparison to healthy cows, which produce an average of 5 liters of milk, diseased cows result in a revenue loss of ZMW 10.00 per cow, highlighting the economic impact of the disease on milk production [17] [19] [20]. Abattoir workers in both study areas reported that animals infected by bTB produce a minimal amount of meat, as some parts, like the lung and the liver, are always rejected [17] [21] [22] [23].

Similarly, study findings also reported that bTB can directly reduce productivity in milk and meat production, particularly in cattle aged 18 months and over [24] [25] [26]. A longitudinal study on the effect of bTB on selected productivity parameters and trading in dairy cattle kept under intensive husbandry in Central Ethiopia found that bTB likely impacts productivity, including lower fertility and weight loss [27] [28] [29]. However, Barnes et al. [20] revealed that only a marginal reduction in reproductive performances and milk yields in bTB-positive animals, contradicting the idea that bTB has a substantial impact on productivity.

Study findings show that males have higher probabilities for a reduction in income than females, attributed to Beef Carcass Condemnation (BCC) [23] [24] [25]. In contrast to BCC, the loss in income due to Low Milk production (LM) shows minimal differences in probabilities between males and females, although females show slightly higher predicted mean probabilities [26] [27] [28]. Men are largely engaged in the business of cattle selling and may be affected by beef and carcass condemnation at an abattoir, hence affecting their financial capabilities [21] [29] [30].

The study findings also confirm that farmers with primary education had their income negatively affected due to low awareness levels about the disease transmission [31][32][33]. Education level significantly influenced farmers’ knowledge and practices regarding bTB in India [22] [34] [35]. Those farmers who were educated were more likely to adopt better management practices and have improved knowledge about the disease.

Respondents aged 40, 50 years and above were highly affected by Bovine Tuberculosis, possibly due to a lack of access to media, low education levels, and limited interactions with veterinary officers [36] [37] [38]. Elderly respondents may have experienced memory decline, affecting their ability to learn and retain new information [39] [40] [41]. Studies also revealed that elderly people are more prone to severe complications from zoonotic diseases due to weakened immunity and underlying health conditions [25] [42] [43] [44] [45].

## CONCLUSION

The study’s findings indicate that Bovine Tuberculosis (bTB) significantly impacts rural livestock farmers in the study areas, hence leading to reduced milk and meat production, increased carcass condemnations, and economic losses. The study, therefore, reveals the relevance of education and awareness in controlling and preventing bTB transmission. It shows that rural elderly people, mostly those with low education levels and limited access to media, are mostly affected by bTB.

### Recommendations

i. There is a need to implement targeted education and awareness programs for rural livestock farmers, particularly focusing on bTB transmission, control, and prevention measures.
ii. There is also a need to maximize open access to veterinary officers and services, particularly in rural areas, to provide regular guidance and support to farmers.
iii. Can come up with effective disease control measures, for instance, as regular testing and vaccination programs, to minimize the prevalence of bTB in the study area.
iv. Intensify comprehensive economic support and compensation to farmers who experience losses due to bTB, to help mitigate the financial impact of the disease.
v. Engage appropriate interventions which will target rural elderly livestock farmer, who are disproportionately affected by bTB, to improve their awareness and knowledge of the disease and its control measures.
vi. Lastly, there is a need to further research the economic impact of bTB in the study area, to inform policy and decision-making, and to develop effective control strategies.

### Limitations

The study findings only focused on bTB; however, other zoonotic diseases or risk factors may also be present in the study area, which could influence the findings. Additionally, the study’s cross-sectional design may not capture the dynamic nature of bTB transmission and its public health impact over time. The study may have underestimated the real impact of bTB on public health because of a lack of awareness about the disease. Lastly, the study was conducted in specific districts, Lundazi and Monze, respectively, which may not be representative of the entire country or region.

## Data Availability

All data produced in the present study are contained in the manuscript

https://orcid.org/0009-0001-5604-8455

## Data availability

The rural livestock farmers provided consent for their data to be used for research and analysis purposes and to publish the findings.

## Acknowledgment

We would like to extend our sincere gratitude to the rural Livestock farmers and communities in Lundazi and Monze districts in the Eastern and Southern provinces of Zambia for participating in this imperative study and sharing their valuable insights, practices, and experiences. Additionally, we would also like to thank the Ministry of Agriculture and Livestock and the Ministry of Health for their support and cooperation.

Lastly, we would like to acknowledge the contributions of our research team, including data collectors and enumerators, for their hard work and dedication.

## Abbreviation Legends

bTB: Bovine tuberculosis
TB: Tuberculosis
WHO: World Health Organization
ZMW: Zambian Kwacha
USD: United States Dollar

